# Clinical performance validation of the STANDARD G6PD Test: A multi-country pooled analysis

**DOI:** 10.1101/2022.12.11.22282391

**Authors:** Wondimagegn Adissu, Marcelo Brito, Eduardo Garbin, Marcela Macedo, Wuelton Monteiro, Sandip Mukherjee, Jane Myburg, Mohammad Shafiul Alam, Germana Bancone, Pooja Bansil, Sampa Pal, Abhijit Sharma, Stephanie Zobrist, Andrew Bryan, Cindy S Chu, Santasabuj Das, Gonzalo J Domingo, Amanda Hann, James Kublin, Marcus VG Lacerda, Mark Layton, Benedikt Ley, Sean C Murphy, Francois Nosten, Dhélio Pereira, Ric N Price, Arunansu Talukdar, Daniel Yilma, Emily Gerth-Guyette

**Author notes:** **Corresponding author:** Emily Gerth-Guyette.

## Abstract

**Introduction:** Screening for G6PD deficiency can inform disease management including malaria. Treatment with the antimalarial drugs primaquine and tafenoquine can be guided by point-of-care testing for G6PD deficiency.

**Methods and Findings:** Data from similar clinical studies evaluating the performance of the STANDARD^™^ G6PD Test (SD Biosensor, South Korea) conducted in Bangladesh, Brazil, Ethiopia, India, Thailand, the United Kingdom, and the United States were pooled. Test performance was assessed in a retrospective analysis on capillary and venous specimens. All study sites used spectrophotometry for reference G6PD testing, and either the HemoCue or complete blood count for reference hemoglobin measurement.

The sensitivity of the STANDARD^™^ G6PD Test using the manufacturer thresholds for G6PD deficient and intermediate cases in capillary specimens from 4212 study participants was 100% (95% Confidence Interval (CI): 97.5%–100%) for G6PD deficient cases with <30% activity and 77% (95% CI 66.8%– 85.4%) for females with intermediate activity between 30%–70%. Specificity was 98.1% (95% CI 97.6%–98.5%) and 92.8% (95% CI 91.6%–93.9%) for G6PD deficient individuals and intermediate females, respectively. The majority (12/20) of G6PD intermediate females with false normal results had activity levels >60% on the reference assay. Negative predictive values for females with G6PD activity >60% was 99.6% (95% CI 99.1%–99.8%) on capillary specimens. Test sensitivity among 396 *P. vivax* malaria cases was 100% (69.2%–100.0%) for both deficient and intermediate cases. In the study population, a high proportion of those classified as G6PD deficient or intermediate resulted from true normal cases. Despite this, the majority cases would receive the correct medication and no true G6PD deficient cases would be treated inappropriately.

**Conclusions:** The STANDARD G6PD Test enables safe access to drugs which are contraindicated for individuals with G6PD deficiency. Operational considerations will inform test uptake in specific settings.

## Introduction

The glucose-6-phosphate dehydrogenase (G6PD) enzyme plays an essential role in the protection of red blood cells against damage from oxidative stress. This enzyme is one of the most polymorphic in the human genome, leading to many mutations resulting in the enzymopathy G6PD deficiency.^1^ G6PD deficiency is one of the most common human genetic disorder affecting an estimated 500 million people worldwide.^1^ The red blood cells of individuals with this condition have decreased G6PD enzyme activity and are more susceptible to hemolysis as a result of an oxidative challenge. Common oxidative challenges include from foods such as fava beans,^2^ infections such as SARS-CoV-2 infection^3,4^ and typhoid, and medications such as rasburicase^5^ and 8-aminoquinoline-based anti-malaria drugs used for radical cure treatment of *Plasmodium vivax* malaria (e.g., primaquine and tafenoquine).^6,7^ G6PD deficiency is also a strong predictor of pathologic neonatal jaundice and potential for life-threatening kernicterus in newborns.^8–10^

Until recently, diagnosis of G6PD deficiency has primarily relied on moderate to high complexity laboratory assays. In practice, the implementation of such tests has been challenging as described in recent publications from the College of American Pathologists.^11,12^ More recently, point-of-care (POC) tests for G6PD deficiency are emerging, providing opportunities to expand testing to populations without access to laboratory-based assays.^13^ Such tests are particularly important in the context of malaria case management, given the limited infrastructure in settings where malaria patients typically seek care. At a global level, the World Health Organization recommends G6PD testing prior to the administration of radical cure treatments for *P. vivax* malaria.^14^ However, among malaria-endemic countries, policies and practices related to G6PD testing and radical cure implementation vary significantly, based in part on the underlying epidemiology of G6PD deficiency as well as barriers to access and adoption.^15^

POC tests for G6PD deficiency include both qualitative rapid tests, as well as quantitative biosensor tests that provide numeric results of patients’ G6PD activity levels. Two qualitative tests have shown promise under laboratory conditions, but have limitations in field conditions, including challenges with temperature correction and the inability to identify females with intermediate G6PD activity arising from a heterozygous *g6pd*_*normal*_*/g6pd*_*deficient*_ genotype.^16–18^ The STANDARD G6PD Test (SD Biosensor, Republic of Korea) is a quantitative enzymatic colorimetric assay intended to aid in the detection of G6PD deficiency at the POC. The test measures G6PD enzyme activity normalized by hemoglobin (Hb) (U/g Hb) and total-hemoglobin concentration (g/dL) on 10 µl of capillary or venous blood samples.

Results are provided within two minutes on a portable, handheld analyzer and used to classify individuals as G6PD normal, intermediate, or deficient according to the manufacturer’s recommended thresholds. This classification can be used to inform clinical decision-making, particularly as it relates to priority applications, such as malaria case management.

Cross-sectional diagnostic accuracy studies have been conducted in a wide range of settings and locations, to evaluate the performance of the STANDARD G6PD Test.^19–22^ Pooling data from these studies allows for a robust diagnostic performance analysis across diverse populations representative of multiple contexts and use cases for POC G6PD testing. Our aim was to 1) present these data for the STANDARD G6PD Test, 2) explore how a common set of thresholds to classify G6PD deficient or intermediate individuals with POC testing can be applied across populations, and 3) consider the implications of the test performance on 8-aminoquinoline malaria treatment regimens.

## Materials and Methods

Data were included from similar cross-sectional diagnostic accuracy studies conducted in seven countries: Bangladesh,^21^ Brazil,^20^ Ethiopia (unpublished), India (unpublished), Thailand,^19^ the United Kingdom,^22^ and the United States^19,22^ (Supplementary Table 1). Collectively, these studies span a varied range of underlying G6PD epidemiology and malaria incidence. The primary objective of all studies was to evaluate the performance of the STANDARD G6PD Test for its ability to identify G6PD normal, intermediate, and deficient individuals, and to evaluate the test’s ability to measure hemoglobin concentration. The performance of the STANDARD G6PD Test on capillary and/or venous specimens was compared to G6PD reference values normalized by hemoglobin from venous specimens tested with a spectrophotometer. Only one study (Thailand) used frozen venous specimens.^19^ The performance of the STANDARD G6PD Test in the measurement of hemoglobin concentration was also compared to a complete blood count from automated hematology analyzers where available, and/or to results from venous specimens on the HemoCue 201+ system.

### Ethical considerations and study populations

All studies involving human subjects were reviewed by relevant ethics committees (Supplementary Table 1) and written informed consent to participate was obtained for all participants. Varying recruitment methods appropriate to each site were employed to ensure study populations that were both representative of intended use settings—including malaria-endemic settings and blood donation centers—as well as of G6PD activity levels across the dynamic range (Supplementary Table 1).

### Testing

#### Supplementary Figure 1 summarizes the tests and overall workflow of the included studies

##### STANDARD G6PD Test

The STANDARD G6PD Test was performed in all studies on non-anticoagulated capillary and/or venous K_2_EDTA whole blood as per the manufacturer’s instructions. In addition to the analyzer, test components include disposable test strips (Test Devices), disposable blood transfer tubes (Ezi Tubes+), extraction buffer vials, and a lot-specific code chip used for calibration. For testing on capillary specimens, 10 μl of blood was collected via fingerprick using a disposable Ezi Tube+ and mixed with the extraction buffer. Next, a second, clean Ezi Tube+ was used to transfer 10 μl of the mixed specimen to the Test Device, which is inserted into the analyzer. When run on venous specimens, μl of blood was transferred to the buffer solution using either a professional pipette or the Ezi Tube+ sample collector.^19,22^ After two minutes, the test reports quantitative measurements of G6PD activity in U/g Hb and hemoglobin (g/dL). The manufacturer’s recommended G6PD activity thresholds were applied to determine G6PD status based on the quantitative test result: 4.0 U/g Hb and 6.0 U/g Hb for G6PD deficient (≤30% activity) and intermediate (≤70% activity), respectively.

##### Reference G6PD testing

In all studies, spectrophotometry was used as the reference assay on venous K_2_EDTA whole blood, using a temperature-regulated instrument. Either the Pointe Scientific (Canton, MI, catalog number G7583) or the Trinity Biotech (Bray, Ireland) G6PD reagent kits were used. All G6PD activity results were normalized for hemoglobin concentration and are presented in U/g Hb.

##### Hemoglobin measurement

Hemoglobin was measured according to the following methods:

- *Complete blood count (CBC).* In the Bangladesh, Brazil, Ethiopia, India, Thailand, and US (2021) studies in Pennsylvania and Washington,^19–22^ total hemoglobin concentration (g/dL) was determined using a CBC on an automated hematology analyzer. This measurement served as the reference method for the measurement of Hb concentration and was used to normalize the reference assay for G6PD activity.
- *HemoCue 201+*. With the exception of the Bangladesh and Thailand studies, the HemoCue Hb 201+ System was also performed on non-anticoagulated capillary blood and/or venous K_2_EDTA blood as a comparative measure of hemoglobin concentration in g/dL according to the manufacturer instructions. For studies where CBC was not available, the HemoCue 201+ measurement was used to normalize the reference assay for G6PD activity.

##### Malaria testing

In three studies, the malaria status of participants was assessed by the standard method at each site—either microscopy (Brazil) or rapid diagnostic test (Ethiopia and India).

### Contrived specimen study

Lastly, data are also presented for a contrived specimen study^22^ that included a panel of 90 specimens spanning critical G6PD activity thresholds developed using heat abrogation and following a method adapted from the UK National External Quality Assurance Services G6PD scheme.^23^ Five contrived specimens representing a broad hemoglobin concentration range were also developed using plasma-level adjustment. These 95 samples underwent blinded testing with the STANDARD G6PD Test and reference assay.

### Statistical methods

Due to the observed inter-laboratory variability of the G6PD reference assay,^11,22^ absolute G6PD values were normalized for each laboratory conducting the reference assay in each study.^24–26^ Reference G6PD activity values were expressed as the percentage of each site’s adjusted male median (AMM). For the majority of studies, the AMM was calculated from a subset of randomly selected males (n=36) with normal G6PD status as determined by each laboratory’s reference range. These 36 males were subsequently excluded from the analytical population for the performance analysis. In the UK study, 39 males were used to calculate the AMM. In the Bangladesh study, the AMM was calculated from the median activity of all known G6PD normal participants. For the Thai study, the AMM was calculated as described in Domingo et al. (2013),^27^ wherein all males with reference G6PD activity ≤10% of the male median were excluded, and a new median activity was determined and used to normalize the reference data. A Kruskal Wallis test was used to compare medians between groups. Next, categorical thresholds were used to classify G6PD deficient cases (males and females with ≤ 30% activity), and females with intermediate activity (> 30% and ≤ 70% activity).

Using pooled data from across all included studies, receiver operating characteristic (ROC) curves were generated to assess the ability of the test to discriminate G6PD deficient males and females, G6PD intermediate females, and G6PD normal males and females.

The pooled performance of the STANDARD G6PD Test against the spectrophotometric reference test for each specimen type was determined by calculating the test’s sensitivity and specificity, and 95% confidence interval (CIs) to diagnose individuals G6PD status at the 30% and 70% thresholds.^27^ For the purposes of this analysis, G6PD deficient and intermediate results by the reference assay were considered as true “ positive”, and G6PD normal results were considered as true “ negatives.” Overall agreement between the STANDARD G6PD Test and reference assay in the classification of normal, intermediate, and deficient G6PD activity levels was calculated and Kappa coefficients were determined at the same thresholds. Agreement was assessed for the STANDARD G6PD Test’s T-Hb measurement against the reference CBC assay results where available, using clinically-relevant thresholds for anemia established by the World Health Organization (WHO) (Supplementary Table 2).^28^

All statistical analyses were performed using Stata® 15.0 (StataCorp, College Station, TX).

To explore the implications of these results for malaria case management, the eligibility of the pooled study population for radical cure treatment regimens was considered. These eligibility criteria were based on the WHO recommendations regarding administration of primaquine for preventing relapse^14^ as well as the product label for tafenoquine (Supplementary Table 3). Across the study population, G6PD classification by the reference assay was compared to G6PD classification by the SD Biosensor test. Next, theoretical treatment eligibility among the study population was calculated based on the results of both the reference assay and the STANDARD G6PD Test, with the former considered as the gold standard and true discriminating test while the latter was considered as the test that would determine treatment status. Based on the concordance or discordance between methods, participants were considered as either correctly or incorrectly receiving or being excluded from the recommended treatment algorithm outcome. Results are presented separately for two different treatment algorithms: 1) primaquine only, and 2) tafenoquine and primaquine, with those who were considered ineligible for tafenoquine then considered for primaquine eligibility. No other treatment eligibility criteria (e.g., age, pregnancy, anemia status) were considered as part of the calculation.

## Results

Data from 4,212 capillary specimens and 4,844 venous specimens were included in the analysis (Table 1). The overall population was split evenly between males and females. The majority of participants with available data had either no or mild anemia. A total of 550 patients with confirmed malaria were included in the analysis.

**Table 1.** Summary of demographics.

|  | Bangladesh<sup>a</sup> | Brazil<sup>b</sup> | Ethiopia | India | UK<sup>c</sup> | US (2021)<sup>c</sup> | Contrived Specimens<sup>c</sup> | US (2019)<sup>d</sup> | Thailand<sup>d</sup> | Total n (%) |
| --- | --- | --- | --- | --- | --- | --- | --- | --- | --- | --- |
| **Specimen type** | Fresh venous K2EDTA | Fresh capillary and venous K2EDTA | Fresh capillary and venous K2EDTA | Fresh capillary and venous K2EDTA | Fresh venous K2EDTA | Fresh capillary and venous K2EDTA | Contrived venous | Fresh venous K2EDTA | Frozen venous K2EDTA |  |
| Final Analytic population, n | 108 | 1,736 | 1,015 | 951 | 167 | 623 | 95 | 174 | 150 | 5,019 |
| Capillary Analytic population, n | N/A | 1,693 | 1,009 | 889 | N/A | 621 | N/A | N/A | N/A | 4,212 |
| Venous Analytic population, n | 108 | 1,662 | 1,015 | 860 | 167 | 613 | 95 | 174 | 150 | 4,844 |
| **Sex, n (%)** |  |  |  |  |  |  |  |  |  |  |
| Female | 81 (75.0) | 948 (54.6) | 476 (46.9) | 345 (36.3) | 87 (52.1) | 309 (49.6) | 90 (97.4) | 73 (41.9) | 108 (72.0) | 2,517 (50.2) |
| Male | 27 (25.0) | 788 (45.4) | 539 (53.1) | 606 (63.7) | 80 (47.9) | 314 (50.4) | 5 (5.3) | 101 (58.1) | 42 (28.0) | 2,502 (49.8) |
| **G6PD status,<sup>e</sup> n (%)** |  |  |  |  |  |  |  |  |  |  |
| Deficient < 30% | 30 (27.8) | 59 (3.4) | 12 (1.2) | 27 (2.8) | 10 (6.0) | 46 (7.4) | 3 (3.2) | 25 (14.4) | 54 (36.0) | 266 (5.3) |
| Intermediate 30%–70% | 21 (19.4) | 35 (2.0) | 19 (1.8) | 17 (1.8) | 10 (6.0) | 17 (2.7) | 58 (61.1) | 10 (5.7) | 46 (30.7) | 233 (4.6) |
| Normal >70% | 57 (52.8) | 1,642 (94.6) | 984 (97.0) | 907 (95.4) | 147 (88.0) | 560 (89.9) | 34 (35.8) | 139 (79.9) | 50 (33.3) | 4,520 (90.1) |
| **Anemia status,<sup>f</sup> n (%)** |  |  |  |  |  |  |  |  |  |  |
| Non/mild | N/A | 831 (47.9) | 473 (46.6) | 360 (37.9) | N/A | 410 (65.8) | 81 (85.3) | N/A | 143 (95.3) | 2,298 (45.8) |
| Moderate | N/A | 83 (4.8) | 1 (0.1) | 91 (9.6) | N/A | 23 (3.7) | 11 (11.6) | N/A | 7 (4.7) | 216 (4.3) |
| Severe | N/A | 10 (0.6) | 0 (0) | 30 (3.2) | N/A | 1 (0.2) | 3 (3.2) | N/A | 0 (0) | 44 (0.9) |
| Missing | 108 (100.0) | 812 (46.8) | 541 (53.3) | 470 (49.4) | 167 (100.0) | 189 (30.3) | 0 (0) | 174 (100.0) | 0 (0) | 2,461 (49.0) |
| **Malaria result,<sup>g</sup> n (%)** |  |  |  |  |  |  |  |  |  |  |
| *P. falciparum* | N/A | 12 (0.7) | 27 (2.7) | 29 (3.1) | N/A | N/A | N/A | N/A | N/A | 68 (1.4) |
| *P. vivax* | N/A | 199 (11.5) | 0 (0.0) | 242 (25.5) | N/A | N/A | N/A | N/A | N/A | 441 (8.8) |
| *P. falciparum/ P. vivax* | N/A | 40 (2.3) | 0 (0.0) | 1 (0.1) | N/A | N/A | N/A | N/A | N/A | 41 (0.8) |
| Negative | N/A | 1,484 (85.5) | 988 (97.3) | 679 (71.4) | N/A | N/A | N/A | N/A | N/A | 3,151 (62.8) |

a. Data published within Alam et al., 2018.
b. Data published within Zobrist et al., 2021.
c. Data published within Pal et al., 2021.
d. Data published within Pal et al., 2019.
e. G6PD status as determined by the spectrophotometric G6PD reference test on venous specimens, based on the specified activity thresholds.
f. Anemia status as measured by CBC in accordance with published clinically relevant Hb concentration thresholds from WHO. The non- and mild anemia categories were combined based on the clinical significance of moderate or severe anemia diagnoses, consistent with the uses of other quantitative devices for the determination of Hb concentration at the point of care. Samples for which only HemoCue Hb 201+ results were available are not included in this summary. Additionally, samples from Bangladesh were not included because of the limitations in the granularity of the data available.
g. Microscopy or rapid diagnostic test positive at recruitment.

### G6PD activity distribution

The site-specific AMMs are reported in Supplementary Table 4. Study-specific median G6PD values and interquartile ranges (IQRs) for all normal males in the analytical populations on both the reference assay and the STANDARD G6PD Test on capillary and venous specimens are presented in Figure 1 and Supplementary Table 5. On the reference assay, the G6PD median ranged from 6.8 U/g Hb to 11.9 U/g Hb, as compared to 6.7 U/g Hb to 11.3 U/g Hb on the STANDARD G6PD Test. Overall, the G6PD median of the STANDARD G6PD Test was within a 1.5 U/g Hb range, with the exception of one of the US studies.^19^ No correlation was observed between the reference assay G6PD normal male median and that of the STANDARD G6PD test across studies (data not shown). In Brazil and India, the study population was recruited from those seeking care for malaria (Supplementary Table 1) and included a sufficient number of *P. vivax* cases among normal males. There was no significant difference in the AMM between malaria negative males and *P. vivax* positive males (Table 2). Among G6PD normal males, the median G6PD activity was significantly lower for *P. vivax* positive males compared to malaria negative males at the Kolkata, India, site, but not at the Porto Velho, Brazil, site.

**Figure 1.**
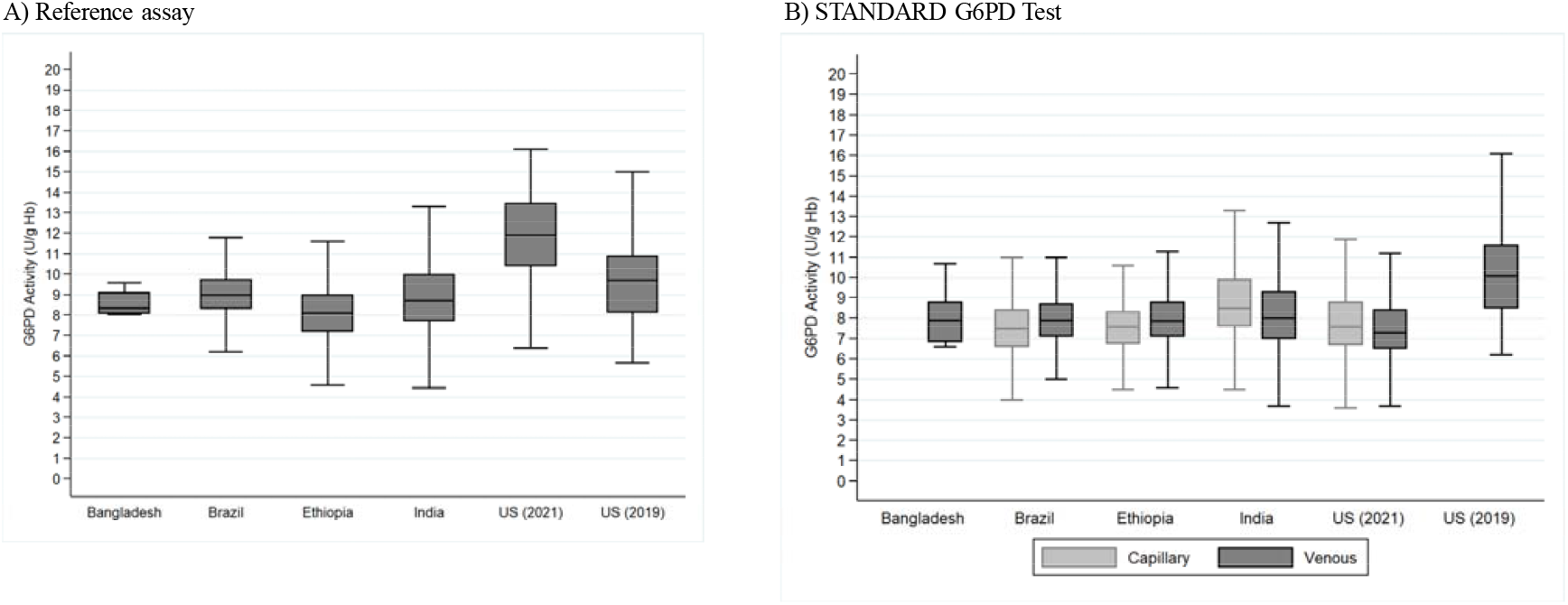
Site-specific variation in G6PD activity among normal males on fresh capillary and venous specimens on A) the reference assay, and B) the STANDARD G6PD Test.* * The UK study, which included newborns and samples with blood disorders, and the Thailand study, which was conducted on frozen samples, are not included in Figure 1.

**Table 2.** Adjusted G6PD normal male median (AMM) and analytical population male medians on the reference assay at the Porto Velho, Brazil, and Kolkata, India, sites, by *P. vivax* malaria status.

|  | Porto Velho, Brazil | Kolkata, India |
| --- | --- | --- |
| AMM | N=36 | N=36 |
| AMM Normal Males<br>Median 100% (95% CI) | 8.9<br>(8.6–9.4) | 8.56<br>(7.9–9.1) |
| AMM *P. vivax* Normal Males<br>Median 100% (95% CI) | 8.7<br>(7.8 – 9.4) | 8.21<br>(7.6 – 9.3) |
| P value | 0.35 | 0.24 |
| All Normal Males (analytical population) | N=368 (all)<br>N=114 (*P. vivax*) | N=582 (all)<br>N=197 (*P. vivax*) |
| All Normal Males<br>Median 100% (95% CI) | 9.1<br>(8.3 – 10.1) | 8.7<br>(7.7 – 10.0) |
| All *P. vivax* Normal Males<br>Median 100% (95% CI) | 8.9<br>(8.1 – 9.8) | 8.2<br>(7.3 – 9.4) |
| P value | 0.08 | 0.001 |

Based on the 30% and 70% activity thresholds, data were available for 266 G6PD deficient, 233 G6PD intermediate, and 4,520 G6PD normal specimens (Table 1). The pooled prevalence for G6PD deficient and intermediate cases in the capillary sample analytical population was 3.4% and 2.1 %, respectively (Supplementary Table 6). The pooled prevalence for G6PD deficient and intermediate cases in the venous blood analytical population was 5.4% and 4.7 %, respectively (Supplementary Table 6). Supplementary Figure 2 presents the distribution of G6PD activities in the study population according to the reference assay and the STANDARD G6PD Test for both males and females.

### Pooled clinical performance of the STANDARD G6PD Test for G6PD

ROC curves (Supplementary Figure 3) and associated performance characteristics (Supplementary Table 7) show that the test performed well in detecting deficient individuals at the 30% activity threshold; the area under the curve (AUC) was 0.998 and 0.997 for capillary and venous specimens, respectively. At the 70% threshold for intermediate females, AUC was 0.909 for capillary and 0.954 for venous specimens.

Table 3 summarizes the pooled diagnostic performance of the STANDARD G6PD Test for identifying G6PD deficient males and females, and females with intermediate G6PD activity, by specimen type. Overall, at the 30% threshold, the test had a sensitivity of 100% for both specimen types. The specificity was 96.7% (95% CI 96.1%–97.2%) for combined venous and contrived specimens and 98.1% (95% CI 97.6%–98.5%) for capillary specimens. For females with intermediate G6PD activity levels, the sensitivity was 77.0% on capillary specimens (95% CI 66.8%–85.4%), 86.6% on venous specimens excluding contrived (95% CI 80.6%–91.3%), and 89.0% on venous specimens including contrived (95% CI 85.4%–93.6%). Specificity was 92.8% for capillary specimens (95% CI 91.6%–93.9%), 94.9% for venous specimens without contrived (95% CI 93.3%–95.3%), and 93.9% for venous specimens including contrived (95% CI 92.8%–94.9%). Overall agreement was 94.8% for capillary and venous specimens, and 93.8% for venous and contrived specimens combined (Supplementary Table 6).

**Table 3.** Summary diagnostic performance of the STANDARD G6PD Test for identifying G6PD deficient males and females and females with intermediate G6PD activity, by specimen type, in all study participants and in *P. vivax* cases only.

| Specimen type | Cases |  |  | Performance for G6PD deficient males and females |  | Performance for G6PD intermediate females (> 30, ≤ 70%) |  | 3 x 3 overall percent agreement (95% CI) | Negative predictive value female > 60% (95% CI) |
| --- | --- | --- | --- | --- | --- | --- | --- | --- | --- |
|  | Deficient cases, n | Intermediate cases, n | Normal cases, n | Sensitivity (95% CI) | Specificity (95% CI) | Sensitivity (95% CI) | Specificity (95% CI) |  |  |
| Capillary | 143 | 79 | 3990 | 100.0 (97.5–100.0) | 98.1 (97.6–98.5) | 77.0 (66.8–85.4) | 92.8 (91.6–93.9) | 94.8 (94.1–95.5) | 99.6 (99.1 – 99.8) |
| Venous (without contrived) | 259 | 110 | 4380 | 100.0 (98.6–100.0) | 97.6 (97.1–98.0) | 86.6 (80.6–91.3) | 94.4 (93.3–95.3) | 94.8 (94.2–95.4) | 99.5 (99.0 – 99.7) |
| Venous (with contrived) | 262 | 152 | 4430 | 100.0 (98.6–100.0) | 96.7 (96.1–97.2) | 89.0 (85.4–93.6) | 93.9 (92.8–94.9) | 93.8 (93.1–94.5) | - |
| ***P. vivax*–infected participants only** |  |  |  |  |  |  |  |  |  |
| Capillary | 10 | 9 | 377 | 100.0 (69.2–100.0) | 99.0 (97.4–99.7) | 100.0 (39.8–100.0) | 93.3 (86.7–97.3) | 96.2 (93.8–97.9) | - |
| Venous | 10 | 4 | 382 | 100.0 (69.2–100.0) | 97.7 (95.6–98.9) | 100.0 (39.8–100.0) | 87.3 (79.2–93.0) | 96.2 (93.8–97.9) | - |

All true deficient individuals were correctly identified by the STANDARD G6PD Test on both venous and capillary specimens and false normal results were only observed for females with intermediate G6PD activity (Figure 2; Table 4). On venous specimens, 10% (23/230) of intermediate females were misclassified as normal, compared to 25% (20/87) on capillary specimen (Table 4). However, the majority (60%, 12/20) of these capillary discordant results were attributed to females with reference G6PD activity greater than 60% and only two samples had G6PD activities between 40 and 50%. (Supplementary Table 8; Table 5; Supplementary Figure 4). Negative predictive values for females with >60% reference activity were 99.6% and 99.5% for capillary and venous specimens (without contrived), respectively (Table 3; Table 5). In contrast, false positive results (particularly for intermediates) occurred in 6% (120/1934) of G6PD normal females who were misclassified as intermediate and 1.5% (34/2178) of G6PD normal males who were misclassified as deficient on capillary specimens.

**Figure 2.**
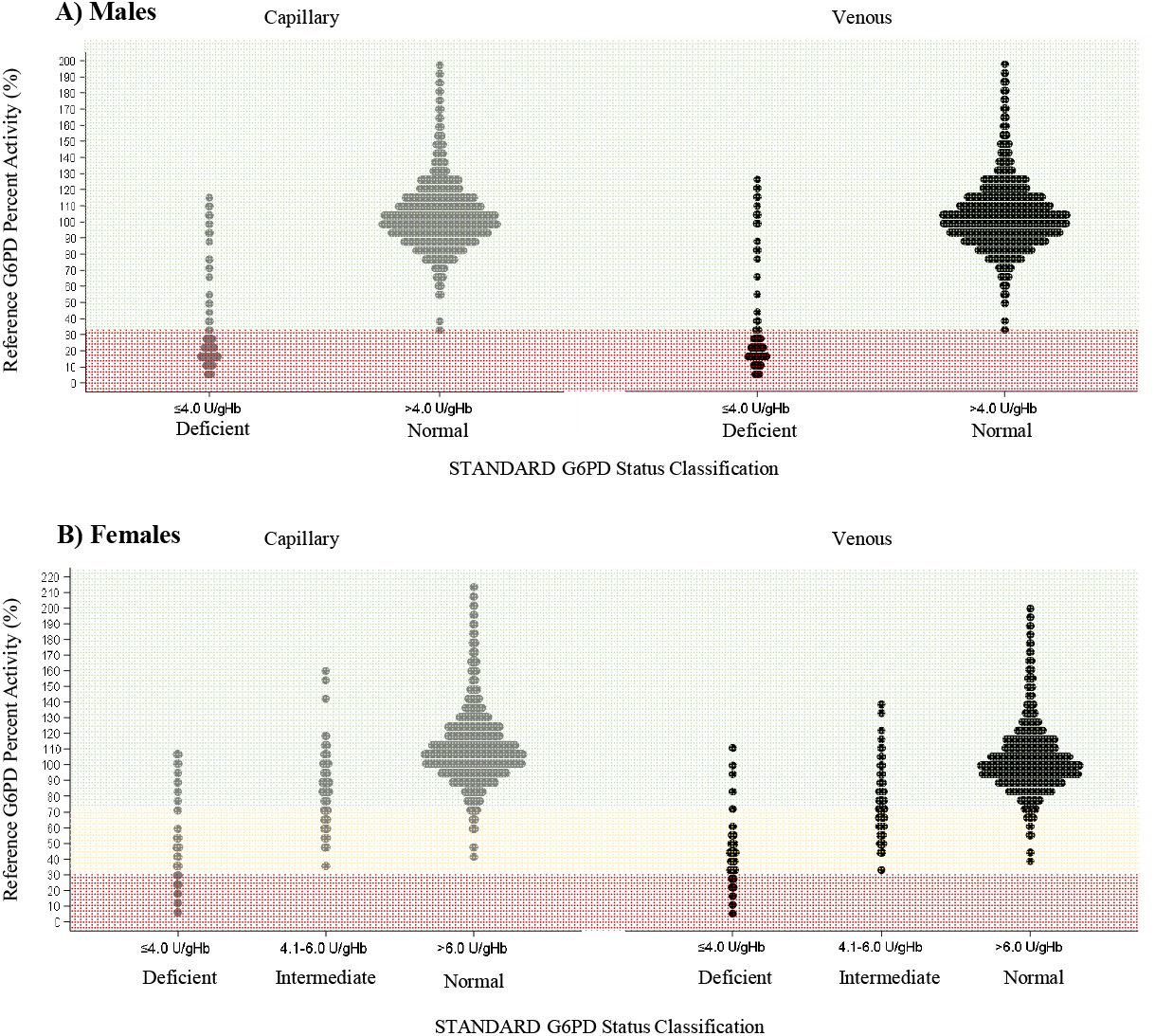
G6PD activity classification of the STANDARD G6PD and reference assays for A) males and B) females. G6PD status classification measured by the STANDARD G6PD test is shown on the X-axis, for capillary (grey) and venous (black) specimens, respectively. Results are plotted by the reference G6PD percent activity (Y-axis). Shaded areas correspond to true G6PD normal (green), intermediate (yellow), and deficient (red) status classifications on the reference assay.

**Table 4.** 2 × 2 and 3 × 3 tables for males and female G6PD classifications respectively by the reference assay (columns) and by the STANDARD G6PD test (rows). 30% and 70% normal G6PD activity thresholds were used to classify G6PD deficient and intermediate status, respectively, with the reference spectrophotometric assay. Manufacturer thresholds on the STANDARD G6PD test were used to classify G6PD deficient and intermediate status.

| STANDARD G6PD<br>Test Classification | Reference G6PD assay classification |  |  | Total |
| --- | --- | --- | --- | --- |
| Capillary: Male |  |  |  |  |
|  | Deficient | Normal |  | Total |
| Deficient | 134 | 30 |  | 164 |
| Normal | 0 | 2018 |  | 2018 |
| Total | 134 | 2048 |  | 2182 |
| Capillary: Female |  |  |  |  |
|  | Deficient | Intermediate | Normal | Total |
| Deficient | 9 | 30 | 19 | 58 |
| Intermediate | 0 | 37 | 120 | 157 |
| Normal | 0 | 20 | 1795 | 1815 |
| Total | 9 | 87 | 1934 | 2030 |
| Venous*: Male |  |  |  |  |
|  | Deficient | Normal |  | Total |
| Deficient | 205 | 34 |  | 239 |
| Normal | 0 | 2144 |  | 2144 |
| Total | 205 | 2178 |  | 2383 |
| Venous*: Female |  |  |  |  |
|  | Deficient | Intermediate | Normal | Total |
| Deficient | 57 | 111 | 7 | 175 |
| Intermediate | 0 | 96 | 125 | 221 |
| Normal | 0 | 23 | 2042 | 2065 |
| Total | 57 | 230 | 2174 | 2461 |

\* Including contrived specimens

**Table 5.** Performance indicators applying the 6.0 U/g Hb threshold on the STANDARD G6PD Test for deficient and intermediate females with G6PD activity levels between 0% and 40%, 50%, 60%, 65%, and 70% for capillary specimens.

|  | % G6PD activity (reference assay) |  |  |  |  |
| --- | --- | --- | --- | --- | --- |
|  | ≤ 70% | ≤ 65% | ≤ 60% | ≤ 50% | ≤ 40% |
| Sensitivity (95% CI) | 79.2<br>(69.7 – 86.8) | 84.6<br>(74.7 – 91.8) | 88.1<br>(77.8 – 94.7) | 92.3<br>(79.1–98.4) | 100.0<br>(83.9–100.0) |
| Number of false normals | 20 | 12 | 8 | 3 | 0 |
| Negative predictive power (95% CI) | 98.9<br>(98.3–99.3) | 99.3<br>(98.8–99.7) | 99.6<br>(99.1–99.8) | 99.8<br>(99.5–100.0) | 100.0<br>(99.8–100.0) |

### Pooled clinical performance of the STANDARD G6PD Test for G6PD among *P. vivax* confirmed cases

Among participants with *P. vivax* malaria, on both capillary and venous specimens, sensitivity was 100% (95% CI 69.2%–100%) at the 30% threshold, with specificities of 99.0% (95% CI 97.4%–99.7%) and 97.7% (95% CI 95.6%–98.9%) for capillary and venous specimens, respectively (Table 3). However, it should be noted that this is based on a small number of deficient and intermediate cases (≤10) within this population.

### Pooled clinical performance of the STANDARD G6PD Test for hemoglobin

Linear regression of the STANDARD G6PD Test’s hemoglobin result as compared to the hemoglobin result on the reference CBC is shown in Supplementary Figure 5, by specimen type. R-squared correlation values are 0.73 and 0.77 for capillary and venous specimens, respectively. Supplementary Table 9 presents the agreement between the STANDARD G6PD Test’s anemia classification on both capillary and venous specimens as compared to the CBC results. Overall percent agreement between methods was 90.3% (95% CI 89.0%–91.5%) for capillary specimens and 94.3% (95% CI 93.2%–95.2%) for venous specimens (excluding contrived). None of the participants with severe anemia (Hb <7.0 g/dL for children 6-59 months of age or <8.0 g/dL for all other groups, n=44) were misclassified as non or mild anemia on either capillary or venous blood.

### Implications for malaria treatment

The STANDARD G6PD test results on capillary specimens were used to calculate the implications of the test performance on eligibility to different radical cure treatment options. Figures 3A and 3B illustrate a scenario where only daily primaquine radical cure regimen is available to males and females with G6PD activity greater than 30%. Based on the results of the STANDARD G6PD test, 94.8% (3993/4212) of the participants, representing all G6PD normal and intermediate participants, would be eligible for standard daily primaquine according to WHO guidelines and correctly provided primaquine based on the concordance between the reference and POC test. Conversely, 5.2% (219/4212) would be excluded from standard primaquine treatment (Figure 3B). No G6PD deficient participants would be prescribed primaquine, but 1.8% (76/4212) males and females with intermediate and normal G6PD activity would be excluded from the daily treatment regimen (Figure 3B; Supplementary Table 10).

**Figure 3.**
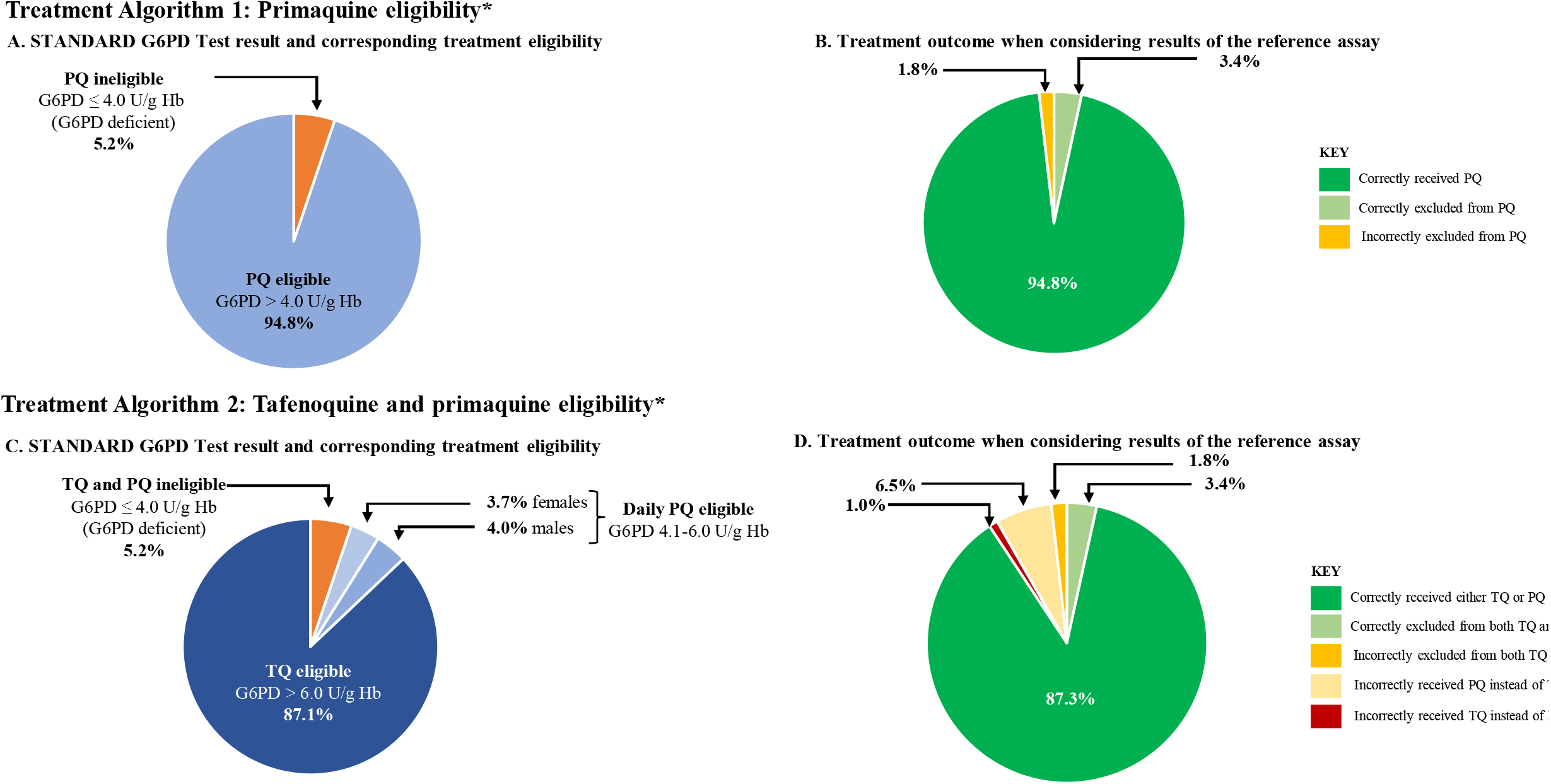
Participant eligibility and outcomes for radical cure treatment options based on the results of the STANDARD G6PD Test on capillary specimens. Panels A and B represent eligibility and outcomes for daily primaquine regimen. Panels C and D represent eligibility and outcomes for tafenoquine and daily primaquine, with those who are considered ineligible for tafenoquine then considered for primaquine eligibility. Abbreviations: G6PD, glucose-6-phosphate-dehydrogenase; PQ, Primaquine; Hb, Hemoglobin; TQ, Tafenoquine * No other treatment eligibility criteria (e.g., age, anemia status, pregnancy status) were considered as part of the calculation.

Figures 3C and 3D illustrate a scenario where both daily primaquine and single dose tafenoquine are available and tafenoquine is only available to males and females with G6PD activity greater than 70%. Based on the POC STANDARD G6PD test results 87.1% (3668/4212) of participants in the study population would be eligible for single dose radical cure (Figure 3C). One percent (43/4212) of the population with G6PD activity ≤ 70% would receive tafenoquine. These were 23 males and 20 females with G6PD activity between 30-70%, with a median G6PD activity of 63%. This analysis found that 7.7% of the population (168 males and 157 females) had a STANDARD G6PD Test result between 4 and 6 U/g Hb and would be eligible for standard daily primaquine all of these had G6PD activity > 30%. Of these, 6.5 % of the total population (153 males and 120 females) had a reference G6PD activity >70% and could have been treated with tafenoquine (Figure 3D; Supplementary Table 10).

## Discussion

In this paper, performance data for the STANDARD G6PD POC test for G6PD deficiency, was consolidated from multiple studies conducted on both capillary and venous specimens across seven countries, representing diverse settings and use cases for G6PD testing.

### Application of universal thresholds

Performance estimates for the STANDARD G6PD Test were calculated by applying a single set of thresholds for G6PD deficiency and female intermediate activity in U/g Hb, as indicated by the manufacturer’s instructions for use. For the reference assay, site-specific adjusted male medians were used to normalize results and establish 30% and 70% G6PD activity thresholds for defining deficient, intermediate, and normal cases. The male median G6PD activity on the POC test was more consistent across sites and specimen types—with the exception of one US site—in comparison to the reference assay, which showed greater site-to-site variability. Inter-site variability in the male median G6PD activity as defined by spectrophotometry has been described previously,^25,29^ and is apparent even between sites testing the same specimens.^22^ Site-to-site reproducibility of the STANDARD G6PD Test conducted on commercially available controls also has decreased variability compared to the reference assay conducted on the same control reagents in the same laboratories.^30^ However, it is important to note with respect to the medians and thresholds that the populations of the studies in this analysis largely comprised of adults and children over the age of two. Only one study (in the United Kingdom) included few newborn samples. Using the STANDARD G6PD Test for newborn screening applications will require the use of higher G6PD threshold values to account for the higher G6PD activity among newborns.^31–34^ A recent study conducted in Thailand with the STANDARD G6PD Test on 307 cord blood samples showed good performance as long as the thresholds are adjusted to address the higher median G6PD activities in newborns.^35^ A recent report suggests that malaria infection may also result in increased G6PD activity when looking across all malaria in deficient and intermediate cases.^36^ However, in our analysis, there was no overall change in G6PD activity when comparing *P. vivax*-infected and uninfected males with normal G6PD activity. The male median G6PD value on the STANDARD G6PD test is statistically consistent across varied contexts and populations (aged two years and older), including in malaria-endemic settings, with the exception of newborns.

### Clinical implications for malaria case management

From a safety perspective, the sensitivity and negative predictive values of POC G6PD tests to identify G6PD deficient and intermediate cases are critical to minimize adverse reactions to drugs that are contraindicated in individuals with G6PD deficiency. This is particularly relevant for the test’s use in malaria case management, as standard 14-day primaquine regimens used for the radical cure of *P. vivax* malaria are not recommended for G6PD deficient males and females,^37,38^ and safety of higher dose regimens among G6PD intermediate females is also a concern.^39^ Additionally, tafenoquine is the first drug to indicate a threshold G6PD activity (70%) on its label, above which data indicates that it is considered safe to prescribe for radical cure.^24,40^ The calculations above suggest that the use of the STANDARD G6PD Test at the point of care reduce risk of drug related hemolysis and so facilitate appropriate provision of treatment, in line with WHO and global treatment guidelines.

The STANDARD G6PD Test showed good performance both on capillary and venous samples for G6PD deficiency when applying the manufacturer’s universal thresholds, and no G6PD deficient cases were misclassified as normal, which ensures G6PD deficient malaria patients will receive correct primaquine regimens. In fact, the STANDARD G6PD Test tends toward overestimation of G6PD deficiency at the deficient and intermediate thresholds as compared to the reference assay (Supplementary Table 6).

The test’s sensitivity decreased for females with intermediate activity. However, most of the false normal results were attributed to females with G6PD activities >60%, (and all of whom had activities >40%), which means that they are less likely to experience severe hemolysis as the potential for severe clinical outcomes increases among those with more severe deficiency (i.e., a low percent G6PD activity). The negative predictive values for females with G6PD reference activity >60% is good at 99.6% and 99.5% for both capillary and venous specimens, respectively. Among the almost 400 *P. vivax* cases included in this analysis, none of the deficient or intermediate cases were incorrectly classified as normal by the STANDARD G6PD Test. However, it should be noted that very few deficient and intermediate *P. vivax*-positive cases were included in this sample.

Of note, challenges associated with false normal G6PD results are not limited to POC tests. For context, a recent report from the College of American Pathologists (CAP) highlights challenges with the accurate diagnosis of specimens with intermediate G6PD activity in current reference clinical laboratory testing.^11^ Among laboratories participating in the CAP proficiency testing, 12.5% of those conducting quantitative reference testing misclassified an intermediate specimen as normal, and 47.8% of those conducting qualitative testing misclassified it as normal.^11^ These findings suggest that misclassifications of G6PD intermediate specimens are not unique to POC tests and that accurate classification of this group is a challenge in clinical laboratory testing.

The specificity of a POC G6PD test is also important when considering performance, as such tests may result in valuable treatment options being withheld from individuals without contraindications. Because of the typically low prevalence of G6PD deficiency in many populations, any decrease in specificity will significantly reduce the positive predictive power of the test with a significant proportion of G6PD deficient and intermediate cases assigned by the POC test actually being G6PD normal individuals. This was observed in the data presented here, where false deficient or intermediate misclassifications on the STANDARD G6PD Test were more common than false normal results. Unpublished data from settings where the STANDARD G6PD test has been scaled up and used outside the context of closely monitored clinical studies indicates that the proportion of false deficient and intermediate cases appears to increase. In the context of malaria, an overestimation of G6PD deficiency may prompt concerns about unnecessarily restricting access to radical cure and higher levels of onward transmission and relapses.

### Implications for malaria programs and future research

Results from this pooled performance analysis will inform future implementation and operational research efforts for this test. One recent usability study conducted among intended test users from three high malaria burden settings found that, with appropriate training, the test can be used in clinics managing malaria cases.^41^ Recent studies also suggest that the incorporation of the test into *P. vivax* case management in Brazil and Laos is cost effective; however, factors such as clinic case burdens and G6PD deficiency prevalence are important considerations.^42,43^ Together, these performance, usability, and cost-effectiveness findings suggest that the STANDARD G6PD Test can be used in malaria endemic settings to support G6PD classification and significantly reduce the risk of drug induced hemolysis when prescribing primaquine or tafenoquine, as well as other drugs such as rasburicase.^44^

However, the positive predictive power of the test among this population, where a significant proportion of individuals classified as G6PD deficient by the STANDARD test are in fact normal represents an operational challenge in settings where confirmatory testing cannot be conducted. As is common for many POC tests, confirmatory testing at a reference laboratory is also recommended if the test indicates a positive result for the disease status.^38,45^ However, where a POC test has the most clinical utility, access to reference confirmatory testing is unlikely. One must consider how to appropriately counsel patients regarding the interpretation of their results.

This and the above limitations in test sensitivity need to be considered in the overall risk benefit assessment of G6PD screening at the point of care, which should also take into consideration local G6PD prevalence and vivax epidemiology as well as other drug eligibility criteria. Additional safety and operational studies may be warranted. Future operational studies should investigate whether testing strategies that involve repeat testing over two or three consecutive occasions can improve confirmation of true G6PD deficient cases and the effective positive predictive value specially in settings with low prevalence in G6PD deficiency. Additionally, given the extremely high negative predictive value (100% for individuals >30% G6PD activity), exploring approaches to retain patients’ G6PD test results could reduce the need for repeat testing for the vast majority of the population, potentially leading to significant cost savings. Finally, the feasibility of using the G6PD test at the community level where malaria care-seeking often takes place will be critical to inform adoption strategies and scale.

### Limitations

There are limitations to this analysis. The different studies contributing to this analysis were not originally designed as a multi-center clinical study, and this pooled analysis was not prospectively planned for all studies. However, the homogeneity in the designs of the included studies with regard to testing methods, data collection, and reporting allows for combining of the data. Importantly, all included studies used one of two compatible reference assays, the Trinity or the Point Scientific spectrophotometric reagent kits, which are based on the same chemistry.^46^ Further, a systematic literature search was not employed to identify included studies. Lastly, our analysis includes fewer data generated from capillary specimens than venous specimens. However, studies that evaluated test performance on capillary specimens included several that were collected from POC settings in malaria endemic countries—Ethiopia, India, and Brazil—and specifically clinics managing malaria cases.

In summary, this pooled analysis supports the use of the STANDARD G6PD Test in near-patient settings where rapid turnaround screening for G6PD status is needed to inform patient care and has previously been unavailable. Operational and costing considerations should inform uptake of the test in specific settings.^47^

## Data Availability

Data from all published studies can be found in the specific publications, as well as in the Harvard Dataverse. Relevant identifiers include the following: Domingo, Gonzalo, 2021, "Fingerstick equivalence and blood disorder interference evaluation of a point-of-care test for G6PD deficiency", https://doi.org/10.7910/DVN/GLLPV9, Harvard Dataverse, V1, UNF:6:KJgifE7Ub32PNxyMZK49Pw== [fileUNF] Zobrist, Stephanie, 2021, "Evaluation of a point-of-care diagnostic to identify glucose-6-phosphate dehydrogenase deficiency in Brazil", https://doi.org/10.7910/DVN/KLUZTX, Harvard Dataverse, V2

https://doi.org/10.7910/DVN/GLLPV9

---

**Supplementary Figure 1.**
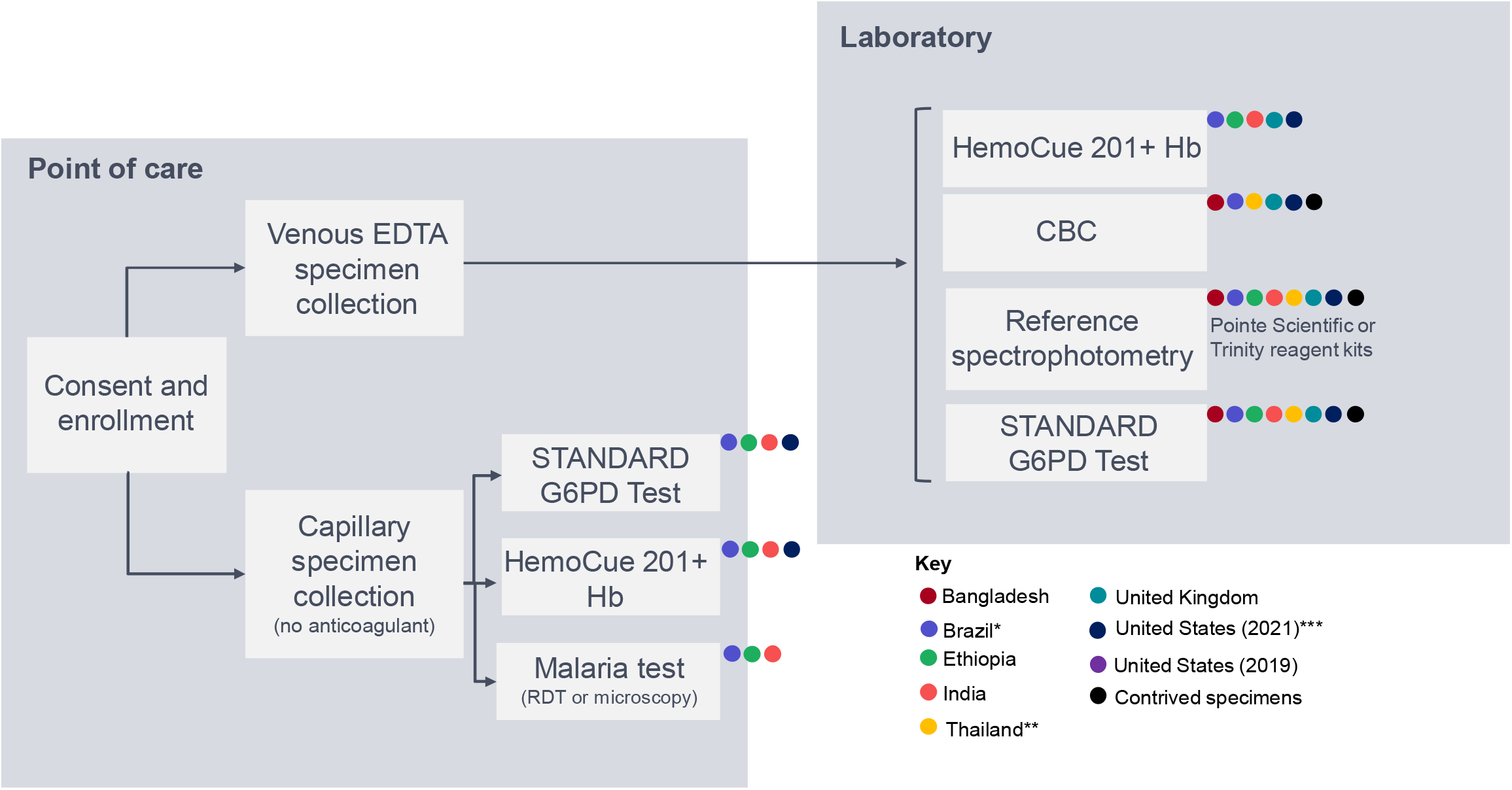
Summary of testing and workflow of included studies. Abbreviations: CBC, complete blood count; FST, fluorescent spot test; G6PD, glucose-6-phosphate dehydrogenase; Hb, hemoglobin

**Supplementary Figure 2.**
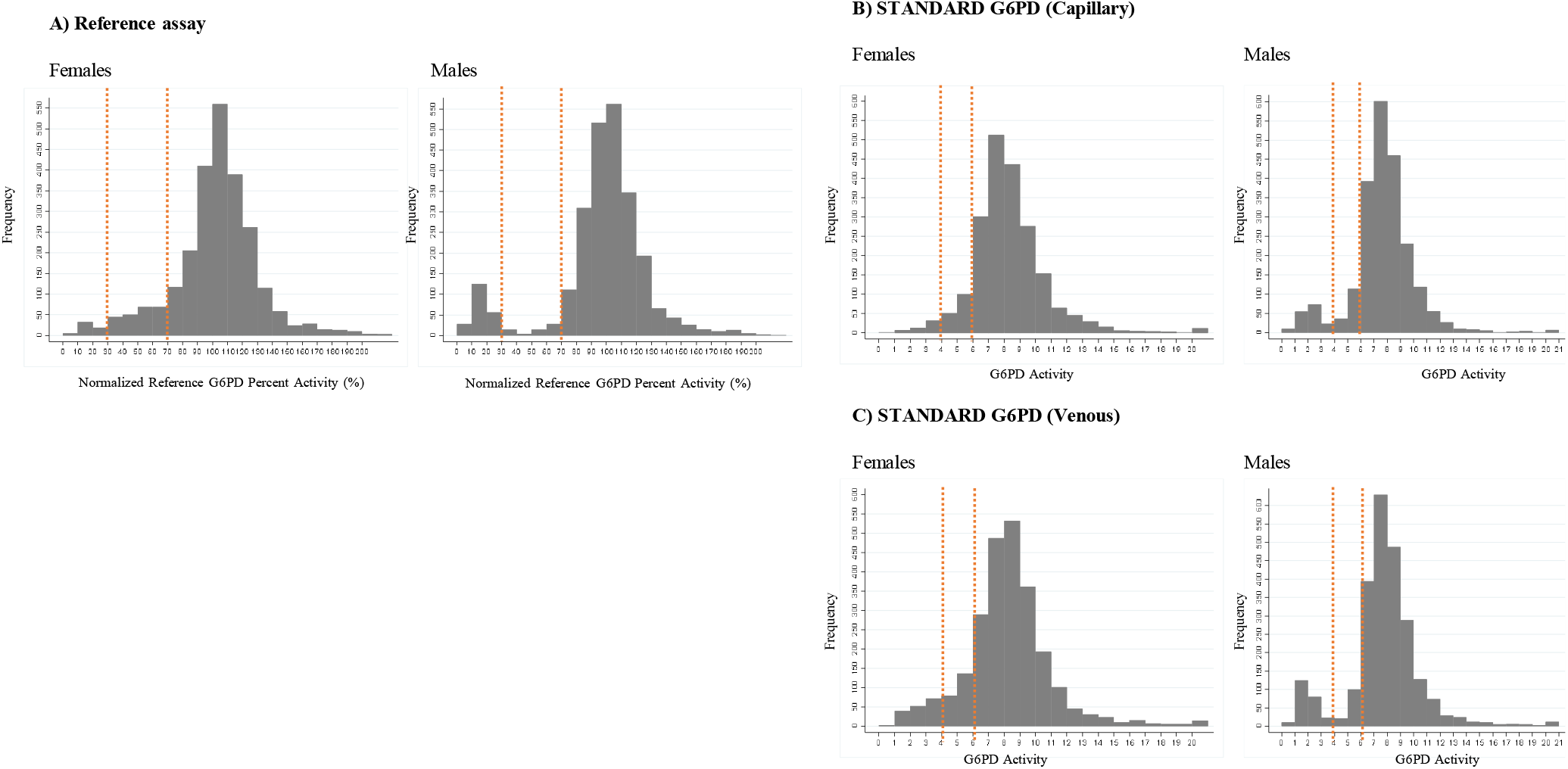
Histograms showing the distribution of G6PD activities in the study population according to A) the reference assay and the STANDARD G6PD Test on B) Capillary and C) Venous specimens. The frequency is plotted in 10% increments for males and females. The 30% and 70% activity limits are indicated on each plot.

**Supplementary Figure 3.**
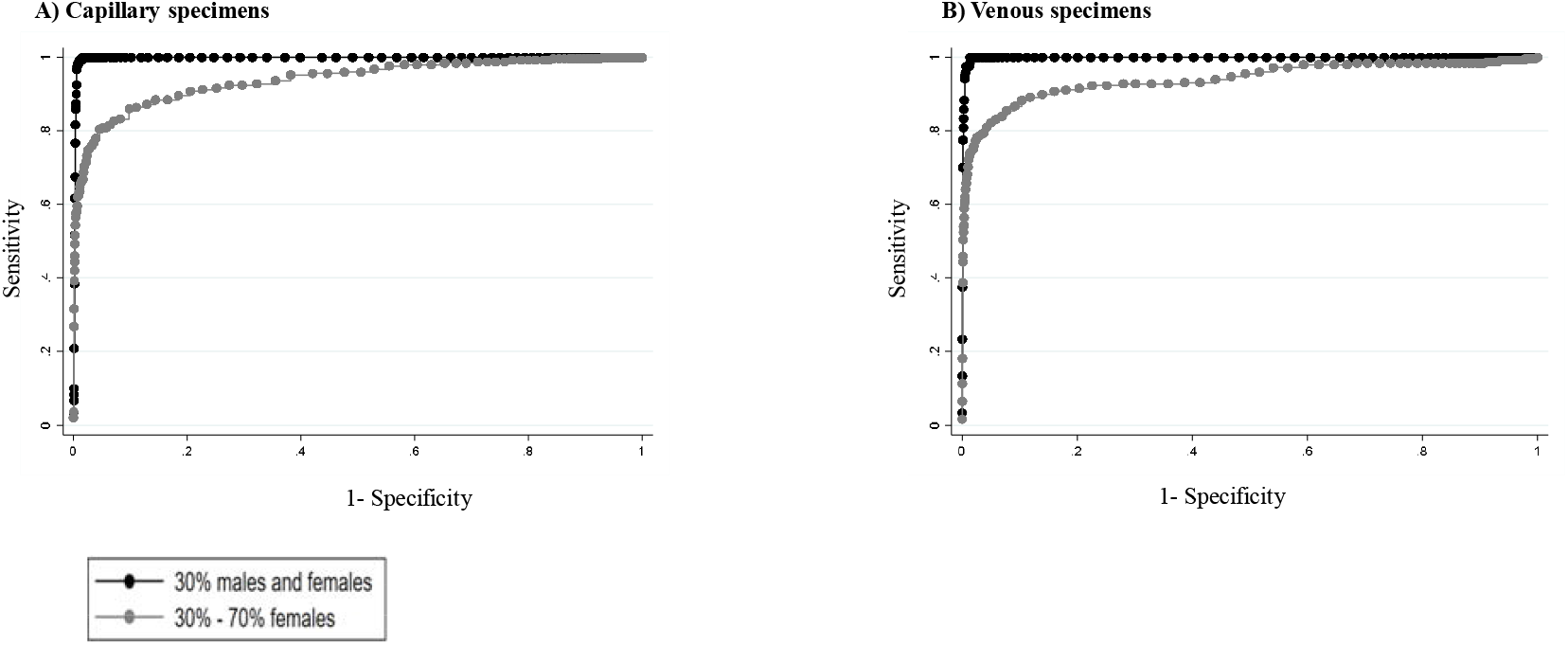
ROC curves at 30% and 70% G6PD activity thresholds on A) capillary specimens and B) venous specimens.

**Supplementary Figure 4.**
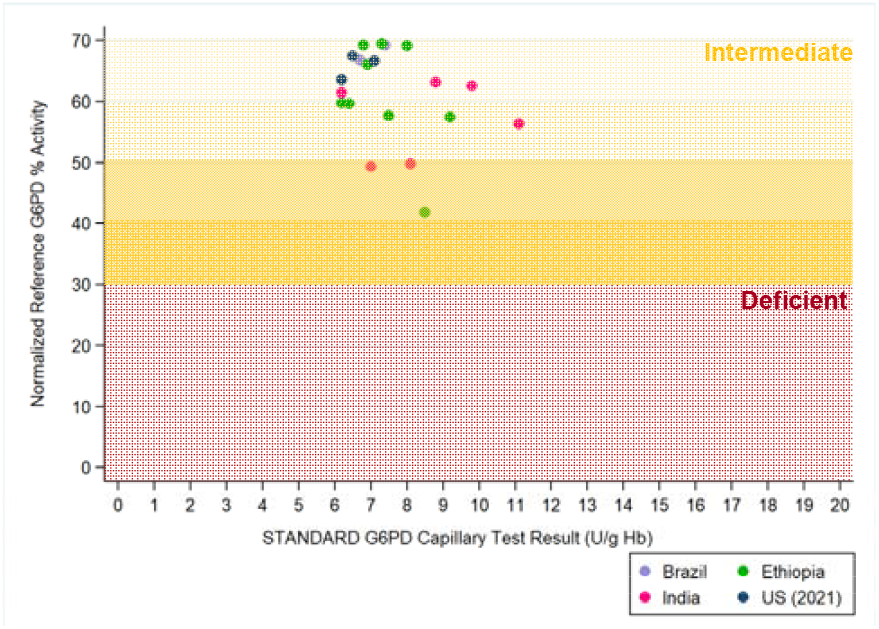
Distribution of false normal capillary results, by study.* * Red shaded areas correspond to deficient (≤30%) reference assay values, and yellow shaded areas correspond to intermediate (30-70%) reference assay values, with darker shading at lower percent activity.

**Supplementary Figure 5.**
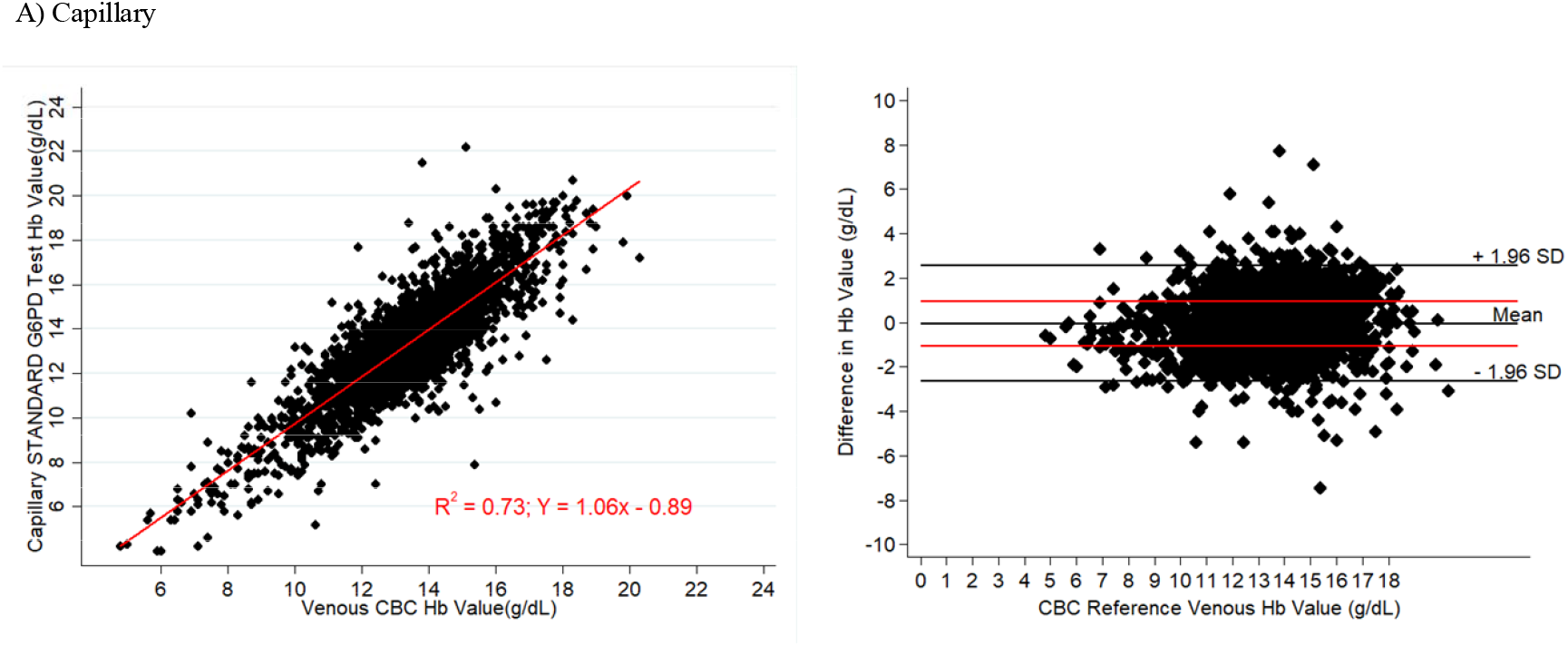
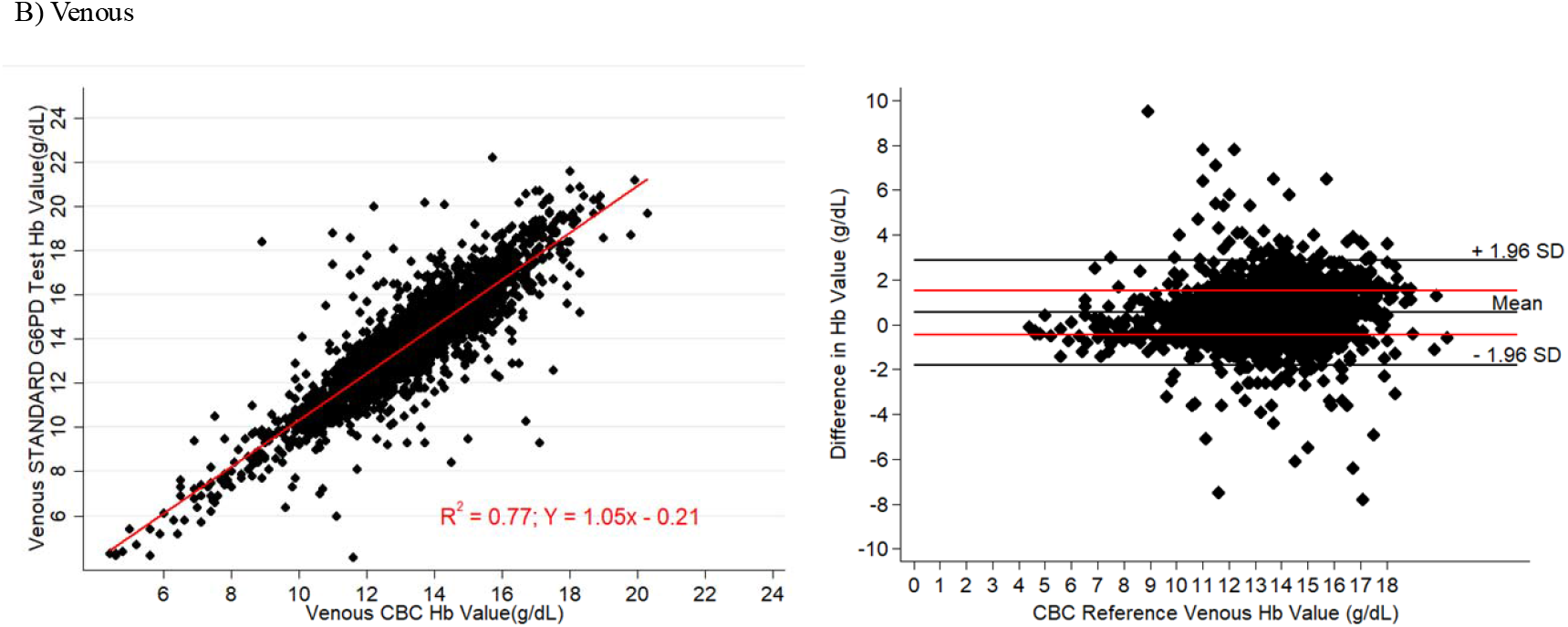
Regression analysis of STANDARD G6PD Test total hemoglobin measurement on A) capillary specimens, and B) venous specimens compared to complete blood count.

