## Supplemental Tables for "Clinical performance validation of the STANDARD G6PD Test: A multi-country pooled analysis"

**Supplementary Materials**

**Supplementary Table 1. Summary of included studies.**

| **Country** | **Number of sites** | **Citation** | **Study type** | **Specimen type** | | | **Study population** | | **Overseeing ethics committee(s)** | **Clinicaltrials.gov identifier, if applicable** |
| --- | --- | --- | --- | --- | --- | --- | --- | --- | --- | --- |
|  |  |  |  | **Fresh capillary** | **Fresh venous K_2_EDTA** | **Frozen venous K_2_EDTA** | **Age** | **Description of sample** |  |  |
| Bangladesh | 1 | Alam et al. 2018 | Prospective |  | X |  | Not available | Individuals with known G6PD status from prior research studies | Ethical review committee (ERC) and research review committee (RRC) of the icddr,b (PR-17043), the Australian Human Research Ethics Committee  (HREC) of the Northern Territory (HREC 17.2771) | N/A |
| Brazil | 2 | Zobrist et al. 2021 | Prospective | X | X |  | ≥2 years | Patients seeking care at clinics in Manaus and Porto Velho, and an enriched sample of participants with known G6PD status from prior research studies | PATH Research Ethics Committee (1204742), Ethics Committee Board of FMT/HVD (94833618.0.1001.0005), Brazil’s  National Research Ethics Commission (CONEP; 94833618.0.1001.0005), CEPEM ethics committee (94833618.0.2001.0011). | NCT04033640 |
| Ethiopia | 2 | Forthcoming | Prospective | X | X |  | ≥2 years | Healthy participants | PATH Research Ethics Committee (1185779)  Ethiopian National Research Ethics Review Committee | N/A |
| India | 2 | Forthcoming | Prospective | X | X |  | ≥8 years | Febrile patients seeking care | PATH Research Ethics Committee (1223628)  Medical College of Kolkata Institutional Ethics Committee,  Indian Council for Medical Research (ICMR)- National Institute of Cholera and Enteric Disease Institutional Ethics Committee | N/A |
| Thailand | 1 | Pal et al. 2019 | Retrospective |  |  | X | ≥18 years | Adults with known G6PD status | Mahidol University Faculty of Tropical Medicine (FTMEC MO/15/259) and the University of Oxford Tropical Research Ethics Committee (OXTREC 563-15) | N/A |
| United Kingdom | 1 | Pal et al. 2021 | Retrospective |  | X |  | ≥2 days | De-identified specimens from completed laboratory testing | Non-research determination from the PATH Research Determination Committee | N/A |
| United States | 3 | Pal et al. 2021 | Prospective | X | X |  | ≥18 years | Healthy adults | Florida: Western Institutional Review Board (20161665)  Pennsylvania: PATH Research Ethics Committee (1416844)  Washington: Fred Hutchinson Cancer Research Center institutional review board [IRB] (10091) | Pennsylvania: NCT04054661  Washington: NCT04010695 |
| United States | 1 | Pal et al. 2019 | Prospective |  | X |  | ≥18 years | African American blood donors from sites in New York and Miami | Western Institutional Review Board (20161665) | N/A |
| Contrived specimen panel | 1 | Pal et al. 2021 | Contrived |  | X |  | N/A | | N/A | N/A |

**Supplementary Table 2. Hemoglobin ranges for anemia (g/dL)**

| **Population** | **Non-anemia** | **Anemia** | | |
| --- | --- | --- | --- | --- |
|  |  | **Mild** | **Moderate** | **Severe** |
| Children 6-59 months of age | ≥ 11 | 10.0–10.9 | 7.0–9.9 | < 7.0 |
| Children 5-11 years of age | ≥ 11.5 | 11.0–11.4 | 8.0–10.9 | < 8.0 |
| Children 12-14 years of age | ≥ 12 | 11.0–11.9 | 8.0–10.9 | < 8.0 |
| Non-pregnant women (15 years of age and above) | ≥ 12 | 11.0–11.9 | 8.0–10.9 | < 8.0 |
| Pregnant women | ≥ 11 | 10.0–10.9 | 7.0–9.9 | < 7.0 |
| Men (15 years of age and above) | ≥ 13 | 11.0–12.9 | 8.0–10.9 | < 8.0 |

Source: World Health Organization. *Haemoglobin Concentrations for the Diagnosis of Anaemia and Assessment of Severity*. World Health Organization; 2011. Accessed March 15, 2021. <https://www.who.int/vmnis/indicators/haemoglobin.pdf>

**Supplementary Table 3. STANDARD G6PD Test result classifications and WHO recommendations for treatment eligibility with Primaquine or Tafenoquine.**

|  | **STANDARD G6PD Test Result** | **G6PD classification or percent activity** | **Recommended or eligible for treatment with primaquine or tafenoquine** |
| --- | --- | --- | --- |
| Primaquine eligibility | Males and females ≤ 4.0 U/g Hb | G6PD deficient | Consider preventing relapse by giving PQ once a week with medical supervision* |
|  | Females 4.1 – 6.0 U/g Hb | G6PD intermediate | Eligible for the 14-day regimen of primaquine, with counselling on how to recognize symptoms and signs of hemolytic anemia |
|  | Males > 4.0 U/g Hb  Females > 6.0 U/g Hb | G6PD normal | Treat in children and adults with a 14-day course in all transmission settings ** |
|  | N/A | G6PD status unknown or testing unavailable | Decision to prescribe PQ based on an assessment of risks and benefits*** |
| Tafenoquine eligibility | Males and females ≤ 6.0 U/g Hb | ≤ 70% | Ineligible for tafenoquine |
|  | Males and females > 6.0 U/g Hb | > 70% | Eligible for tafenoquine **** |
|  | N/A | G6PD status unknown or testing unavailable | Ineligible for tafenoquine |

**Note, this regimen is not commonly prescribed in practice and for the purposes of this analysis, those testing as G6PD deficient are considered to not have access to PQ or TQ*

***Except pregnant women, infants aged < 6 months, women breastfeeding infants aged < 6 months, women breastfeeding older infants unless they are known not to be G6PD deficient and people with G6PD deficiency.*

**** Risks include low relapse rates, low P. vivax incidence rates, high G6PD deficiency prevalence, patient unable to detect signs and symptoms of hemolysis, and patient has poor access to health care system*

***** Eligibility is also currently restricted to those older than 16 years of age, though pediatric studies are ongoing*

Source: World Health Organization (WHO). Guide to G6PD deficiency rapid diagnostic testing to support P. vivax radical cure. Published online 2018. Licence: CC BY-NC-SA 3.0 IGO.

**Supplementary Table 4. Site-specific adjusted male medians (AMMs) used for reference-assay threshold determination.**

|  | **Bangladesh^a^** | **Brazil^b^** | | **Ethiopia** | | **India** | **UK^c^** | **US (2021) ^c^** | | **US (2019)^d^** | **Thailand^d^** |
| --- | --- | --- | --- | --- | --- | --- | --- | --- | --- | --- | --- |
|  |  | **Manaus** | **Porto Velho** | **Jimma** | **Gambella** |  |  | **Pennsylvania, Washington, and Contrived specimens** | **Florida** |  |  |
| Median 100% | 9.9 | 8.63 | 8.93 | 8.11 | 7.89 | 8.56 | 8.7 | 12.85 | 7.49 | 9.03 | 6.84 |
| 30% | 2.97 | 2.58 | 2.67 | 2.43 | 2.37 | 2.57 | 2.61 | 3.87 | 2.25 | 2.71 | 2.05 |
| 70% | 6.93 | 6.02 | 6.23 | 5.67 | 5.52 | 5.99 | 6.09 | 9.03 | 5.24 | 6.32 | 4.79 |

a. Data published within Alam et al., 2018.

b .Data published within Zobrist et al., 2021.

c. Data published within Pal et al., 2021.

d. Data published within Pal et al., 2019.

**Supplementary Table 5. Median G6PD values and interquartile ranges from all normal males in the analytical populations*, by site***

|  | **Bangladesh^a^** | **Brazil^b^** | **Ethiopia** | **India** | **UK ^c^** | **US (2021)^c^** | **US (2019)^d^** | **Thailand^d^** |
| --- | --- | --- | --- | --- | --- | --- | --- | --- |
| G6PD median (IQR) Pointe Scientific, venous specimens  (U/g Hb) | 8.3  (8.1– 9.1) | 9.0  (8.3 – 9.7) | 8.1  (7.2 – 9.0) | 8.7  (7.7 -10.0) | 11.5  (10.2 -14.2) | 11.9  (10.4 – 13.5) | 9.7  (8.1 – 10.9) | 6.8  (6.6 – 7.8) |
| G6PD median (IQR) STANDARD G6PD Test, capillary specimens (U/g Hb) | N/A | 7.5  (6.6 – 8.4) | 7.6  (6.8– 8.3) | 8.5  (7.6 – 9.9) | N/A | 7.6  (6.7 – 8.8) | N/A | N/A |
| G6PD median (IQR) STANDARD G6PD Test, venous specimens (U/g Hb) | 7.9  (6.9 – 8.8) | 7.9  (7.1 – 8.8) | 7.9  (7.1 – 8.8) | 8.1  (7.1 – 9.5) | 11.3  (9.4 – 13.9) | 7.3  (6.5 – 8.4) | 10.1  (8.5 – 11.6) | 6.7  (5.7 – 7.7) |

Abbreviation: IQR, interquartile range.

* Where applicable, excluding data from normal males that were used to calculate the adjusted male medians (AMM).

a. Data published within Alam et al., 2018.

b .Data published within Zobrist et al., 2021.

c. Data published within Pal et al., 2021.

d. Data published within Pal et al., 2019.

**Supplementary Table 6. Percent agreement between the STANDARD G6PD Test and the reference assay using the manufacturer’s threshold values at 30% and 70% G6PD activity thresholds, on A) capillary specimens, B) venous specimens (excluding contrived), and C) Venous specimens (including contrived).**

A. Capillary

|  | | **G6PD cases defined by the reference assay** | | | **Total** |
| --- | --- | --- | --- | --- | --- |
|  |  | **Deficient** | **Intermediate** | **Normal** |  |
| **STANDARD G6PD Test** | **Deficient** | 143 | 30 | 49 | 222 |
|  | **Intermediate** | 0 | 37 | 120 | 157 |
|  | **Normal** | 0 | 20 | 3,813 | 3,833 |
|  | **Total** | 143 | 87 | 3,982 | 4,212 |

Percent agreement between hemoglobin-normalized G6PD activity categorized results and the STANDARD G6PD Test was 94.8% [95% CI: 94.1–95.5].

Kappa: 0.69

B. Venous (excluding contrived)

|  | | **G6PD cases defined by the reference assay** | | | **Total** |
| --- | --- | --- | --- | --- | --- |
|  |  | **Deficient** | **Intermediate** | **Normal** |  |
| **STANDARD G6PD Test** | **Deficient** | 259 | 69 | 41 | 369 |
|  | **Intermediate** | 0 | 80 | 113 | 193 |
|  | **Normal** | 0 | 23 | 4,164 | 4,187 |
|  | **Total** | 259 | 172 | 4,318 | 4,749 |

Percent agreement between hemoglobin-normalized G6PD activity categorized results and the STANDARD G6PD Test was 94.8% [95% CI: 94.2–95.4].

Kappa: 0.69

C. Venous (including contrived)

|  | | **G6PD cases defined by the reference assay** | | | **Total** |
| --- | --- | --- | --- | --- | --- |
|  |  | **Deficient** | **Intermediate** | **Normal** |  |
| **STANDARD G6PD Test** | **Deficient** | 262 | 111 | 41 | 414 |
|  | **Intermediate** | 0 | 96 | 125 | 221 |
|  | **Normal** | 0 | 23 | 4,186 | 4,209 |
|  | **Total** | 262 | 230 | 4,352 | 4,844 |

Percent agreement between hemoglobin-normalized G6PD activity categorized results and the STANDARD G6PD Test was 93.8% [95% CI: 93.1–94.5].

Kappa: 0.69

**Supplementary Table 7. Areas under the curve (AUC) for receiver operating characteristics (ROC) analysis of the performance of the STANDARD G6PD Test for G6PD activity against the reference test for G6PD-deficient males and females as well as intermediate females, by specimen type.**

| **STANDARD G6PD activity** | **AUC** |
| --- | --- |
| **Capillary specimens** | |
| 30% activity males and females | 0.998 |
| 70% activity females only | 0.909 |
| **Venous specimens** | |
| 30% activity males and females | 0.997 |
| 70% activity females only | 0.954 |

**Supplementary Table 8. Summary of discordant false negative results on A) Capillary specimens and B) Venous specimens*.**

A. Capillary

| **Participant identifier** | **Site** | **Reference % G6PD activity** | **Capillary STANDARD G6PD Test G6PD Result (U/g Hb)** |
| --- | --- | --- | --- |
| ETGA-1037 | Ethiopia | 0.42 | 8.5 |
| INCC-0256 | India | 0.49 | 7 |
| INMC-0504 | India | 0.50 | 8.1 |
| INMC-0505 | India | 0.56 | 11.1 |
| ETGA-1220 | Ethiopia | 0.57 | 9.2 |
| ETGA-1034 | Ethiopia | 0.58 | 7.5 |
| ETGA-1285 | Ethiopia | 0.60 | 6.4 |
| ETGA-0981 | Ethiopia | 0.60 | 6.2 |
| INMC-0490 | India | 0.61 | 6.2 |
| INMC-0509 | India | 0.63 | 9.8 |
| INCC-0409 | India | 0.63 | 8.8 |
| USPA159 | US^c^ | 0.64 | 6.2 |
| ETGA-1331 | Ethiopia | 0.66 | 6.9 |
| G243 | US^c^ | 0.67 | 7.1 |
| BRMA-0667 | Brazil^b^ | 0.67 | 6.7 |
| USPA206 | US^c^ | 0.67 | 6.5 |
| ETGA-1221 | Ethiopia | 0.69 | 8 |
| BRMA-0015 | Brazil^b^ | 0.69 | 7.4 |
| ETGA-0985 | Ethiopia | 0.69 | 6.8 |
| ETGA-1289 | Ethiopia | 0.69 | 7.3 |

B. Venous*

| **Participant identifier** | **Site** | **Reference % G6PD activity** | **Venous STANDARD G6PD Test G6PD Result (U/g Hb)** |
| --- | --- | --- | --- |
| ETGA-1037 | Ethiopia | 0.42 | 8.20 |
| INCC-0256 | India | 0.49 | 8.20 |
| INMC-0504 | India | 0.50 | 8.80 |
| ETGA-0905 | Ethiopia | 0.55 | 6.10 |
| INMC-0505 | India | 0.56 | 13.40 |
| BRH1420534 | US^d^ | 0.57 | 7.00 |
| ETGA-1220 | Ethiopia | 0.57 | 8.40 |
| B82 | Bangladesh^a^ | 0.58 | 6.90 |
| ETGA-1034 | Ethiopia | 0.58 | 8.40 |
| ETGA-1285 | Ethiopia | 0.60 | 6.60 |
| ETGA-0981 | Ethiopia | 0.60 | 6.40 |
| INMC-0490 | India | 0.61 | 6.30 |
| INMC-0509 | India | 0.63 | 11.10 |
| INCC-0409 | India | 0.63 | 7.60 |
| USPA159 | US^c^ | 0.64 | 6.40 |
| B141 | Bangladesh^a^ | 0.67 | 8.70 |
| BRH1420533 | US^d^ | 0.67 | 8.70 |
| ETGA-1221 | Ethiopia | 0.69 | 7.80 |
| ETGA-0985 | Ethiopia | 0.69 | 7.20 |
| ETGA-1289 | Ethiopia | 0.69 | 6.20 |
| BRMA-0415 | Brazil^b^ | 0.70 | 6.40 |

* Data from Thailand study (Pal et al., 2019) has not been included.

a. Data published within Alam et al., 2018.

b .Data published within Zobrist et al., 2021.

c. Data published within Pal et al., 2021.

d. Data published within Pal et al., 2019.

**Supplementary Table 9. Percent agreement tables for anemia status between the STANDARD G6PD Test and the reference Complete Blood Count (CBC) T-Hb measurement for a) Capillary specimens and b) Venous specimens (excluding contrived)**

A. Capillary

|  | | **CBC** | | | |
| --- | --- | --- | --- | --- | --- |
|  |  | **Severe anemia** | **Moderate anemia** | **Non/mild anemia** | **Total** |
| **STANDARD**  **G6PD Test** | **Severe anemia** | 33 | 32 | 2 | 66 |
|  | **Moderate anemia** | 3 | 127 | 149 | 279 |
|  | **Non/mild anemia** | 0 | 33 | 1,888 | 1,921 |
|  | **Total** | 36 | 192 | 2,039 | 2,267 |

Percent agreement between CBC and the STANDARD G6PD Test was 90.3% [95% CI: 89.0–91.5].

B. Venous (excluding contrived)

|  | | **CBC** | | | |
| --- | --- | --- | --- | --- | --- |
|  |  | **Severe anemia** | **Moderate anemia** | **Non/mild anemia** | **Total** |
| **STANDARD**  **G6PD Test** | **Severe anemia** | 33 | 11 | 2 | 46 |
|  | **Moderate anemia** | 4 | 135 | 69 | 208 |
|  | **Non/mild anemia** | 0 | 49 | 2,051 | 2,100 |
|  | **Total** | 37 | 195 | 2,122 | 2,354 |

Percent agreement between CBC and the STANDARD G6PD Test was 94.3% [95% CI: 93.2–95.2].

**Supplementary Table 10. 3x3 agreement tables between the STANDARD G6PD Test and the reference assay G6PD percent activity, on capillary specimens for A. males, and B. females.**

**A. Males**

|  | | **Reference assay % activity** | | | **TOTAL** |
| --- | --- | --- | --- | --- | --- |
|  |  | **≤30%** | **30-70%** | **>70%** |  |
| **STANDARD G6PD Test** | ≥6.1 U/g Hb | 0 | 23 | 1830 | 1853 |
|  | 6-4 U/g Hb | 0 | 15 | 153 | 168 |
|  | ≤ 4 U/g Hb | 134 | 11 | 16 | 161 |
|  | **TOTAL** | 134 | 49 | 1999 | 2182 |

**B. Females**

|  | | **Reference assay % activity** | | | **TOTAL** |
| --- | --- | --- | --- | --- | --- |
|  |  | **≤30%** | **30-70%** | **>70%** |  |
| **STANDARD G6PD Test** | ≥6.1 U/g Hb | 9 | 30 | 19 | 58 |
|  | 6-4 U/g Hb | 0 | 37 | 120 | 157 |
|  | ≤ 4 U/g Hb | 0 | 20 | 1795 | 1815 |
|  | **TOTAL** | 9 | 87 | 1934 | 2030 |
